# Assessment of the effectiveness of required weekly COVID-19 surveillance antigen testing at a university

**DOI:** 10.1101/2023.11.22.23298917

**Authors:** Christopher W. Ryan

**Author notes:** Corresponding author. Address: SUNY Upstate Medical University Binghamton Clinical Campus, PO Box 778, Johnson City, NY, 13790, US.

## Abstract

**Objectives:** To mitigate the COVID-19 pandemic, many institutions implemented a regimen of periodic required testing, irrespective of symptoms. The effectiveness of this”surveillance testing” requires assessment.

**Methods:** I fit a zero-inflated negative binomial model to COVID-19 testing and case investigation data between 1 November 2020 and 15 May 2021, from young adult subjects in one community. I compared the duration of symptoms at time of specimen collection in those diagnosed via (1) surveillance testing at a university, (2) the same university’s student health services, and (3) all other testing venues.

**Results:** The data comprised 2926 records: 393 from surveillance testing, 493 from student health service, and 2040 from other venues. About 65% of people with COVID-19 detected via surveillance testing were already symptomatic at time of specimen collection.

Predicted mean duration of pre-testing symptoms was 1.7 days (95% CI 1.59 to 1.84) for the community, 1.81 days (95% CI 1.62 to 1.99) for surveillance, and 2 days (95% CI 1.83 to 2.16) for student health service. The modelled “inflated” proportions of asymptomatic subjects from the surveillance stream and the other/community stream were comparable (odds ratio 0.95, p = 0.7709). Comparing surveillance testing with the student health service, the proportion of “excess” zero symptom durations was signficantly higher in the former (Chi-square = 12.08, p = 0.0005)

**Conclusions:** Surveillance testing at a university detected 393 people with COVID-19, but no earlier in their trajectory than similar-aged people detected in the broader community. This casts some doubt on the public health value of such programs, which tend to be labor-intensive and expensive.

**Three-question summary box:** *What is the current understanding of this subject?:* Assessments of long-term operational effectiveness of COVID-19 “surveillance testing” have not been published.

*What does this report add to the literature?:* During the 2020-2021 academic year at one university, people with COVID-19 detected via compulsory weekly surveillance antigen testing were equally likely to be symptomatic at time of detection, and for just as long, as similar-aged people detected via testing venues in the community.

*What are the implications for public health practice?:* Surveillance testing programs during the pandemic consumed a large amount of time, money, and effort. In future respiratory pandemics, resources might be better devoted to other mitigation measures.

## Introduction

Hoping to control the spread of COVID-19 attributable to asymptomatic and presymptomatic infections, many institutions implemented a regimen of periodic required testing, irrespective of symptoms, for people notionally under their jurisdiction—often called “surveillance testing.” Among them were many colleges and universities. After abruptly halting in-person, on-campus instruction in March 2020, surveillance testing was widely considered important for a safe return to in-person collegiate education in fall 2020 and was widely adopted.^1–12^

Early mathematical modeling studies attempted to predict the effects of surveillance testing,^2,13–19^ and several pilot or short-duration studies of implementation were also conducted.^4,6,7^ As these programs are costly and labor-intensive, it is now important to assess their effects from a longer-term perspective. The present study concerns a single university (“University”) where, serendipitously, surveillance test results could be confidently distinguished from those generated by the student health service and by other testing venues in the wider community. The research question is whether University’s COVID-19 surveillance testing program detected COVID-19 infections signficantly earlier in their clinical evolution, compared to University’s own student health servece, and to the broader community where the the like-aged population was not generally subject to surveillance testing.

## Methods

The analysis involved all positive COVID-19 test results and the subsequent case investigation reports among subjects aged 17.5 to 27 years, inclusive, in a county of approximately 198,000 population, between 1 November 2020 and 15 May 2021. For most of that interval, wild-type SARS-CoV-2 was the predominant circulating strain; by the end of the study period, the alpha strain accounted for about half of sequenced specimens worldwide.^20^ The county encompasses University, which enrolls approximately 18,000 students, undergraduate and graduate. Generally healthy people of study age became eligible for COVID-19 vaccination toward the end of the study period.^21^ The Institutional Review Board of SUNY Upstate Medical University determined that this project comprised public health surveillance and did not meet the definition of human subjects research (determination 1933720-1).

During the study period, the local health department was obligated to investigate all occurrences of COVID-19 via structured telephone interview, which included the presence and duration of symptoms. All those with COVID-19 received official isolation orders and instructions. Exposed contacts disclosed by the patients were also interviewed and quarantined. The health department maintained continuous, close coordination with University.

Weekly, students at University were required to have a BINAXNow COVID-19 antigen test,^22^ conducted at the student union (“surveillance testing”). This began in late August, but with a different testing product; that period is excluded from this analysis. Students with symptoms of COVID-19 were not to attend their next scheduled surveillance test but rather seek clinical evaluation promptly. For many, this would be via University’s student health service. Nevertheless, many students disclosed symptoms at the surveillance site, and if their test was positive, they were automatically considered by the local health department to have COVID-19. Subjects with a positive antigen test but who denied symptoms at the surveillance site were sent, immediately or the next day, to the student health center for a point-of-care Cue Health isothermal nucleic acid amplification test (NAAT).^23^ If positive on NAAT, they too were considered to have COVID-19 and investigated as above; if negative (discordant results), they were not considered infected and no further action was taken.

Employees of University were also subject to weekly required surveillance testing and are subsumed here under the term”students.” There is no reliable, automated way to distinguish laboratory results pertaining to students from those pertaining to employees.

Ill students presenting to the student health center were tested as clinically indicated, with one or more of an antigen test, a point-of-care NAAT, or a specimen sent to a reference laboratory for a NAAT (“shs testing”). Employees were not generally eligible for clinical care at the student health center, except for the follow-up NAAT for those asymptomatic but with a positive surveillance test.

Students could also avail themselves of other testing venues: one state-operated and two hospital-operated drive-through NAAT sites, and a mobile antigen testing service operated by the local health department. These venues are here aggregated under the category “community/other testing.”

The results of all COVID-19 tests were reported into the state’s Electronic Clinical Laboratory Reporting System (ECLRS) and thence to the local health department where the person resides. College students attending University were considered residents of the study county, although in practice some results were mis-routed to the health department of their family home; those were not accessible for this study. I used all COVID-19 test results present in the study county’s ECLRS database as of 14 February 2023.

Each result record contained a Clinical Laboratory Improvement Act (CLIA) number that uniquely identified the laboratory that analyzed the specimen. By convenient coincidence, University used different CLIA numbers for its surveillance testing and its student health service testing, a decision for made for internal, operational reasons. All non-University CLIA numbers were categorized here as “community/other.”

All positive COVID-19 laboratory reports were automatically transmitted into a commercial database called CommCare (Dimagi, Inc.), which was provided by the state for this one purpose. CommCare served as the data system for guiding the investigator’s interview and for recording the findings. I used all records of investigations present in CommCare as of 27 December 2022.

The lab result and investigation datasets were each separately filtered to eliminate those with a variety of anomalies, including:

- specimen collection date prior to February 2020 or as dates that had not yet occurred
- duplicate entries: results from the same specimen from the same person on the same day
- investigations marked as entered in error, duplicate, or outside the local health department’s jurisdiction

Using the fastLink package^24^ in R,^25^ I then matched the two data sources on the basis of last name, first name, day of the month of the birthdate (1-31), and age in fractional years on the date the specimen was collected. Records were considered successfully matched if the posterior probability of an accurate match was greater than 85%.

I then filtered the matched records to those meeting the following criteria:

- specimen collection date (SCD) was within the study period
- the laboratory stream was recorded unambiguously
- the presence or absence of symptoms was recorded unambiguously
- if symptoms were present, the symptom onset date (SOD) was within seven days either side of the SCD
- the SCD in ECLRS was within four days either side of the SCD in CommCare

This yielded the final, analytical dataset.

Using the countreg package^26–28^ in R, I fit a zero-inflated negative binomial model, with the number of pre-test symptomatic days as the response and the testing stream by which the infection was detected as the predictor of interest. The “other/community” stream served as the baseline category. When symptoms began after specimen collection or were entirely absent, the response variable was set to zero. All students with COVID-19 first detected via a positive surveillance antigen test were attributed to that testing stream, irrespective of any immediate follow-up NAAT testing at student health service. I controlled for overall community incidence with a kernel smoothed estimate of the proportion of tests across the entire county (all three testing streams) that were positive each week.

Briefly, a zero-inflated negative binomial model accomodates a mixture distribution where subjects with zero counts (here, zero pre-collection symptomatic days) can arise in two different ways: via a count process with overdispersion, or via a dichotomous process in which a count is either zero or non-zero. A zero-inflated model can accomodate a larger number of subjects with zero counts than can be well-fit by typical Poisson or negative binomial models—a so-called “excess” of zeros.

I assessed goodness of model fit with graphical methods in the countreg pacakge. I compared coefficient estimates on the lab streams with Wald tests. Lastly, I conducted 5000 simulations from the model, at a typical age of 22 years and at two different levels of overall community incidence, generating point estimates and confidence intervals for stream-specific probability distributions of the number of pre-test symptomatic days.

## Results

The county-wide percent of tests each week that were positive ranged from 1.14% to 12%, with a median of 5.52%.

The query of the state laboratory database yielded 2292320 records. Excluding those with anomalous dates left 1384786. Limiting those to subjects aged 17.5 to 27 years left 340051, of which 12887 were positive.

The query of the Commcare case investigation database yielded 97543 records. Excluding records marked as duplicate, erroneous, or out-of-jurisdiction left 76615. Of those, 8935 were between 17.5 and 27 years of age, inclusive. All non-erroneous records in Commcare by definition represent patients with positive test results.

Record linkage yielded 7870 matched records. After the exclusions itemized above and summarized in Table 1, 2926 matched records remained. These comprise the final, analytical dataset to which models were fit.

**Table 1:** Reasons matched records were excluded from further analysis and modeling. SOD = symptom onset date. SCD = specimen collection date. Reasons are not mutually exclusive, i.e. some records may have qualified for exclusion on more than one account. Total number excluded was 4944

| exclusion criterion | number excluded |
| --- | --- |
| more then 4 days discordance on SCD | 670 |
| missing indicator of symptoms | 175 |
| ambiguous lab indicator | 88 |
| more than 7 days between SCD and SOD | 662 |
| missing duration between SCD and SOD | 1986 |
| prior to study period | 628 |
| after study period | 3950 |

The testing streams from which the 2926 records in the analytical dataset emerged are summarized in Table 2.

**Table 2:** Sources of positive COVID-19 test results in the final analytical dataset: other = non-university-affiliated settings; shs = university student health service; surveillance = university surveillance testing.

| testing stream | number | percent |
| --- | --- | --- |
| other | 2040 | 70% |
| shs | 493 | 17% |
| surveillance | 393 | 13% |

The distribution of elapsed days between SOD (as determined from the case investigation interview) and SCD (as determined from the state laboratory result database) is shown in Figure 1.

**Figure 1:**
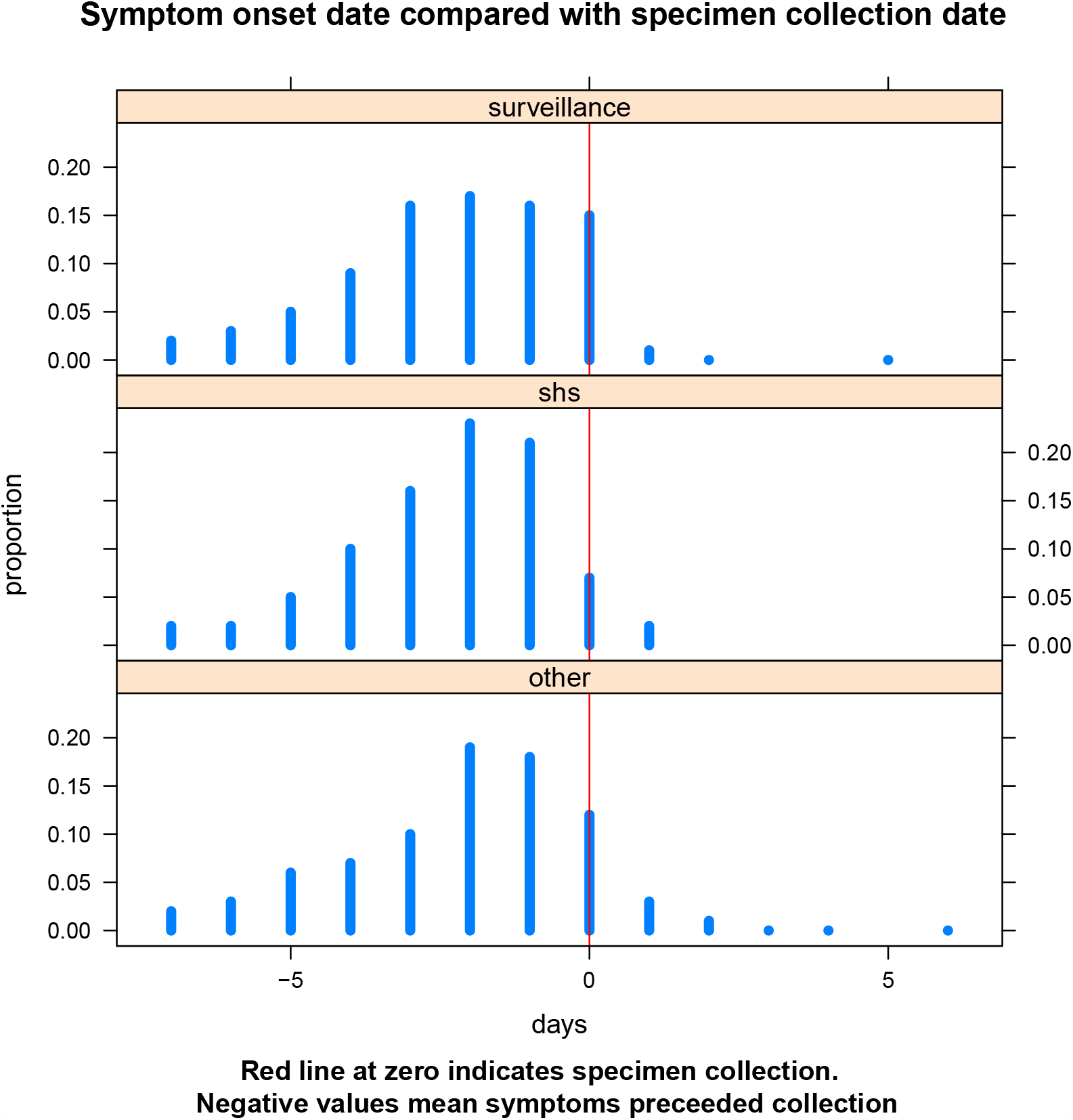
Distribution of times between onset of symptoms (from the case investigation interview) and the specimen collection date (from the state laboratory result database) in the three different testing streams, as a proportion of all results from the respective streams. Negative values mean onset of symptoms preceded specimen collection; conversely, positive values mean onset of symptoms followed specimen collection.

### Modeling results

The fitted model is shown in Table 3. The coefficient estimates and standard errors in Table 3 are on the scale of the linear predictor, so positve values imply an increase in the probability of an “excess” zero (top half of Table 3) or in the number of pre-collection symptomatic days discounting the excess zeros (bottom half of Table 3). Model fit seems adequate, as explored in more detail in the Supplemental Materials.

**Table 3:**
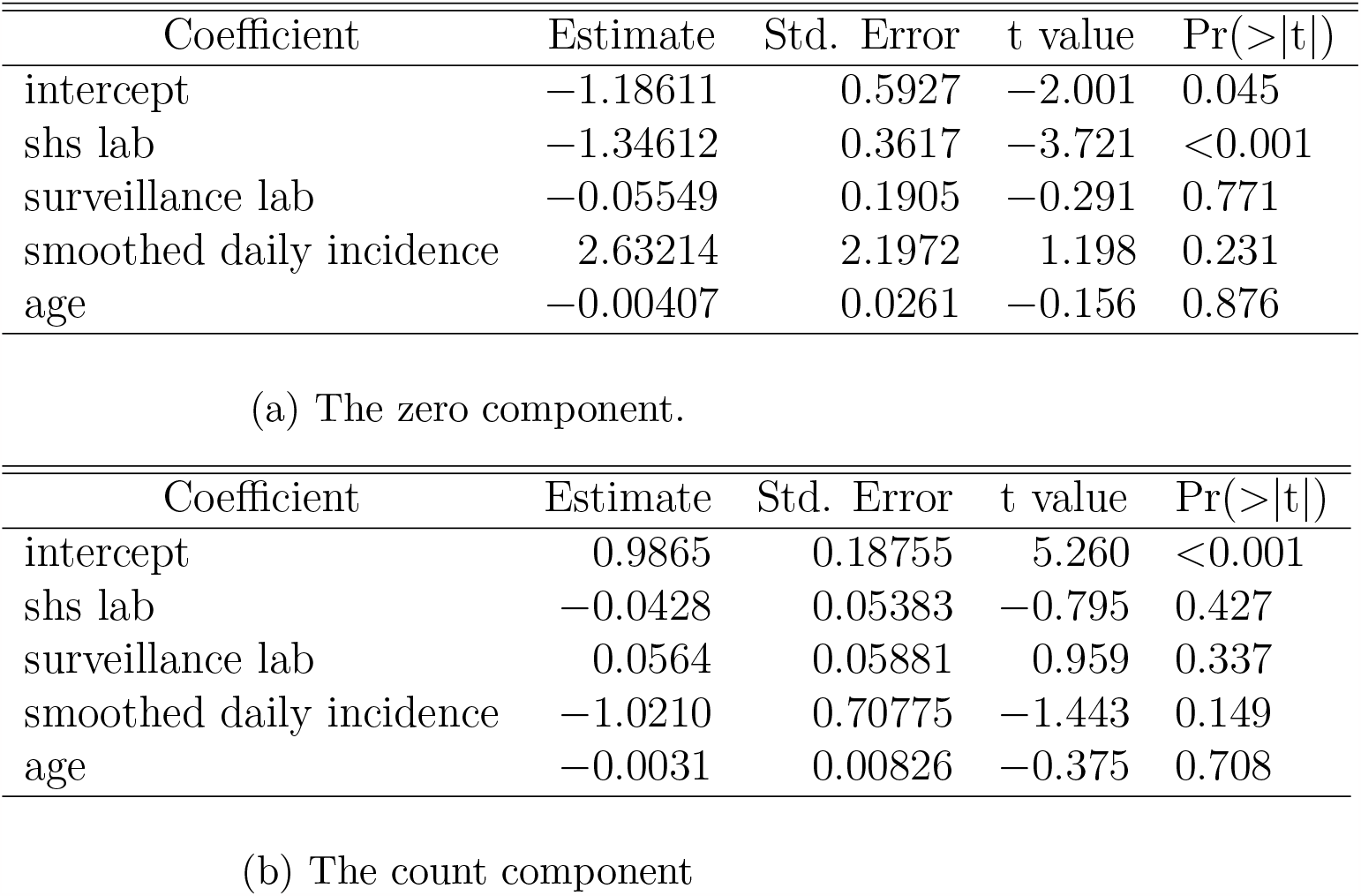
Zero-inflated negative binomial model. The coefficient estimates and standard errors are on the scale of the linear predictor. The “other” lab stream, i.e. the broader community outside University, serves as the baseline category.

Part 3a of Table 3 models the “excess” portion of subjects asymptomatic at time of specimen collection. These “excess zeros” are those poorly accounted for by the negative binomial count process modeled in subtable 3b. Subtable 3a suggests that the “excess” proportion of asymptomatic subjects presenting to the student health service (shs) is significantly lower than that in the other/community setting (odds ratio 0.26, p = 0.0002.) This is perhaps not surprising. On the other hand, the “excess” proportions of asymptomatic subjects from the surveillance stream and the other/community stream were comparable (odds ratio 0.95, p = 0.7709). Comparing surveillance testing with the student health service, the proportion of “excess” zero symptom durations was signficantly higher in the former (Chi-square = 12.08, p = 0.0005)—also not surprising.

Part 3b of Table 3 models the negative binomial count process for the number of symptomatic days prior to specimen collection (which can itself also yield zeros). Subtable 3b suggests that, discounting the “excess” asymptomatic infections (that were accounted for by the “excess zero” component in 3a), the mean duration of pre-collection symptoms was not significantly different in either the surveillance stream or the student health stream, compared to the baseline other/community stream (odds ratios 0.96, p = 0.4265 and 1.06, p = 0.3374, respectively.)

Combining the two components of the model, we can predict two informative things:

1. The probability that a person detected as having COVID-19 would have any particular number of pre-test symptomatic days. This is shown in Figure 2, for each lab stream, at each of two illustrative overall community incidence levels: “low” (the observed lower quartile of 3.9%) and “high” (the observed upper quartile of 8.75%). At either incidence level, the probability profiles of University’s surveillance testing stream and that from the wider community are nearly identical. On the other hand, people identified via the student health service stream were less likely to have been asymptomatic.

**Figure 2:**
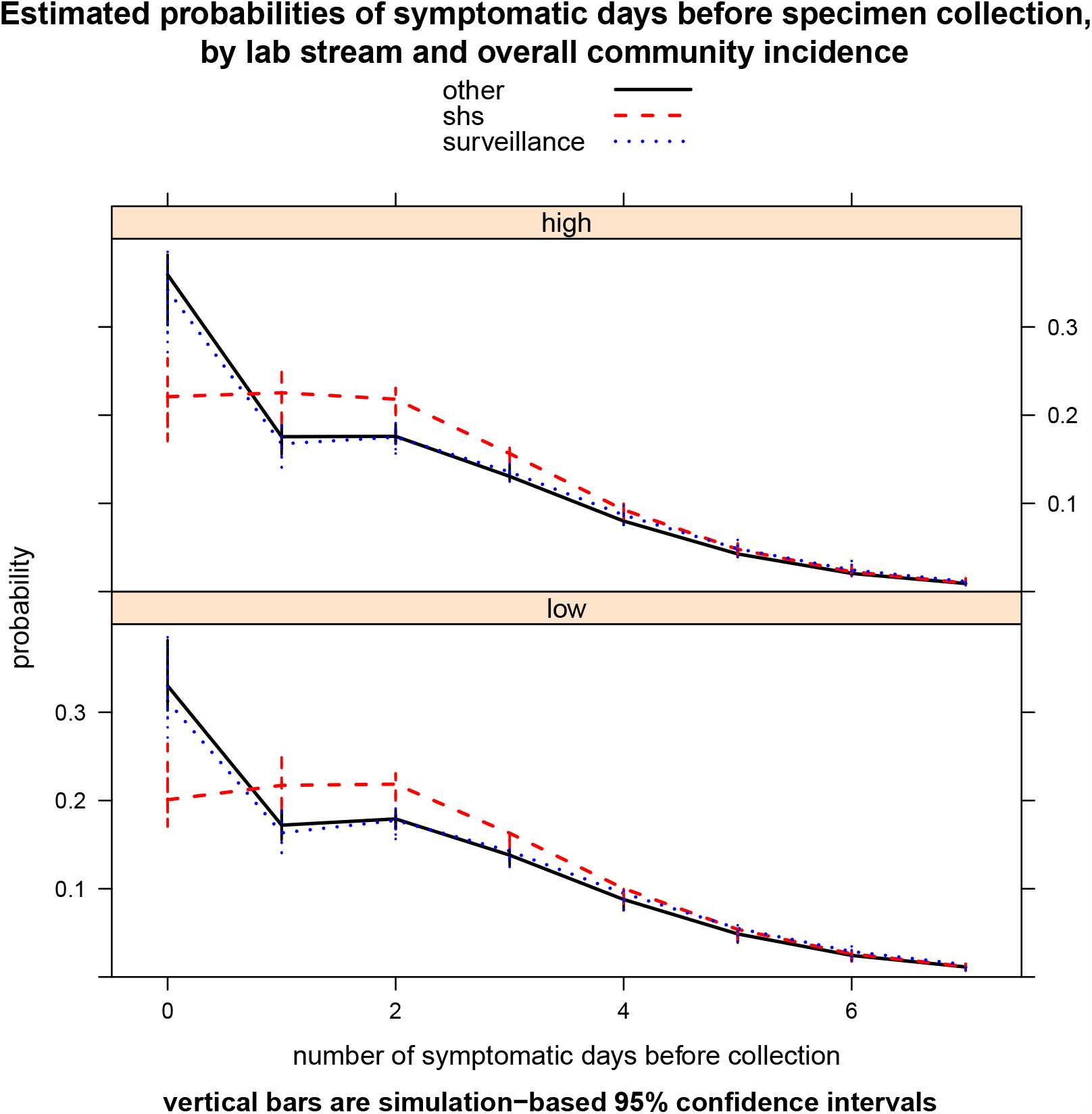
The model-estimated proportion of subjects with any given number of symptomatic days (from 0 to 7) prior to specimen collection, at each of two levels of community incidence, for each of three laboratory streams. The probability of symptom duration beyond 7 days was very low and is not shown. “shs” = student health service.
2. The mean duration of symptoms prior to specimen collection. This is shown in Figure 3 for each lab stream, again at “low” and “high” community incidence. Within each graph, there is much overlap between predicted mean pre-collection symptom durations from each of the three testing streams. The scale of the horizontal axis should also be noted: what differences there may be in the predicted means are on the order of a few tenths of a day, i.e. 6-12 hours.

**Figure 3:**
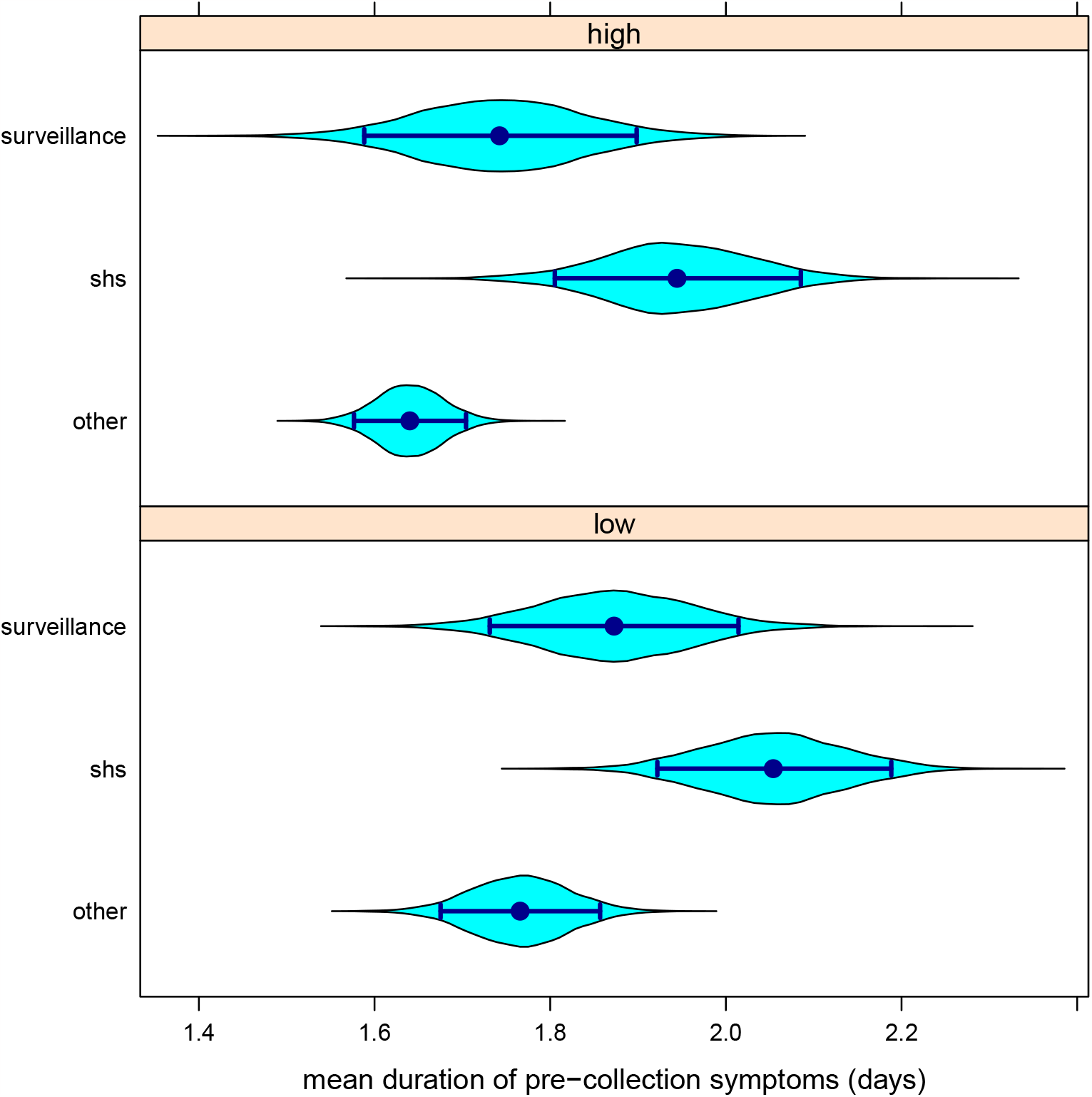
5000 values of mean duration of symptoms simulated from the model, at each of two levels of community incidence, for each of three laboratory streams. Envelopes represent the density of the mean duration values obtained, while the dark bars show the means and their 95% two-sided confidence intervals. “shs” = student health service.

## Discussion

At a three-year remove from the onset of the pandemic, the effects of surveillance testing programs can now be assessed with some perspective. The serendipitous use of a separate CLIA number for surveillance testing at University enabled the present analysis to compare surveillance testing with (1) University’s student health service and (2) testing venues in the wider community used by college-aged people.

The interpretation of the present findings is nuanced. About 35% of students with COVID-19 detected via University’s surveillance regimen had no symptoms at the time of specimen collection.

This compares with about 20% of those detected via the student health service. This could be considered a success for disease control, in that it enabled early isolation of those asymptomatic but infected students, and identification and quarantine of their contacts.

Nevertheless, that leaves about 65% of surveillance-detected students already symptomatic at the time of detection, despite instructions that symptomatic students should not wait for their scheduled surveillance test. The concern is that those students could potentially transmit infection while waiting. On the other hand, a more encouraging interpretation is that those infections might have gone entirely undetected but for University’s surveillance regimen.

The fundamental question in the present analysis is whether infections with COVID-19 were detected earlier in their trajectory among a population subject to surveillance testing compared to a like-aged population that generally was not. This analysis provided no evidence that they were. Those identified via University’s surveillance testing were as likely to be symptomatic at the time of specimen collection as similar-aged subjects arising from various testing venues in the community, and model-predicted mean duration of pre-collection symptoms differed only trivially between the three different testing streams. These findings raise important questions about the utility of resource-intensive surveillance testing programs in mitigating respiratory pandemics.

Public health interventions take place in a larger behavioral, social, ecological, economic, occupational, and academic context. These contextual factors may influence the effects of such programs, perhaps by influencing care-seeking (or care-avoiding) behavior—in potentially paradoxical ways.

In any one-month period, approximately 75% of adults will develop some sort of symptom, yet only one-third of those will seek medical care.^29–31^ Symptoms are by definition subjective and not verifiable externally. They also have meaning for the person experiencing them, and that meaning can change with time and events. Minor symptoms initially self-interpreted as unworthy of mention or action may later be reinterpreted, especially after being presented with a test result.^32^

People with COVID-19 could not attend class, work, or other group activities. Neither could the people who were identified as having been close around them. So besides the medical ramifications, the decision to seek prompt clinical testing, as opposed to awaiting a scheduled surveillance test, entailed opportunity costs, and not just for the person infected.

“Risk homeostasis,” still a controversial concept, posits that people might become less careful when they perceive their individual risk to be reduced by external mitigation measures.^33,34^ This could manifest as a delay in seeking care if one felt reassured by the results of a recent (negative) surveillance test or by the sure knowledge of another test coming up soon. Several early mixed-methods studies in university settings suggested that some subjects receiving a negative result on a recent test indeed felt reassured and thus willing to liberalize their social interactions.^35–39^

## Limitations

This analysis was based on data from weekly antigen-based surveillance testing at a single university and may not be generalizable. Replication in other settings, where other procedures were used, would be valuable.

Release from isolation depended mainly on the presence and duration of symptoms, which were measured by telephone interview after diagnosis. It is possible that some subjects fabricated symptoms, or provided spuriously early symptom onset dates, to expedite their release. This caveat would apply to both students (subject to surveillance testing) and non-students (not generally subject to surveillance testing), so any bias may not have been differential.

A small number of county residents unaffiliated with University were subject to surveillance testing, mainly the employees of skilled nursing facilities. Test results from these settings were included here in the “other/community” stream, since they could not easily be otherwise distinguished. Compared to University, these surveillance testing programs were relatively low-volume, with a still smaller fraction of participants expected to be of study age.

No subjects were interviewed specifically for the purposes of this study. The case investigation records did not record any subject’s thoughts, feelings, or intentions around getting or not getting tested through any particular stream. That would be fertile ground for further study.

## Conclusions

People with COVID-19 detected via a compulsory weekly COVID-19 surveillance antigen testing program at a university were equally likely to be symptomatic at time of detection, and for just as long, as similar-aged people whose COVID-19 was detected via testing venues in the broader community. This casts some doubt on the public health value of such programs, which are generally expensive and labor-intensive. Given the mean duration of symptoms found here, a twice- or thrice-weekly testing frequency may have been necessary to achieve ealier detection, with a concomitant increase in the effort and resources required. Alternatively, in future respiratory pandemics, resources might be better devoted to other mitigation measures.

## Supporting information

Assessment of Model Fit

## Data Availability

Original data are not available to others due to confidentiality concerns.

## References

[1] N. J. Matheson et al. “Mass testing of university students for covid-19.” In: BMJ (Clinical research ed.) 375 (Oct. 2021), 2388. issn: 1756-1833. doi: 10.1136/bmj.n2388.epublish.

[2] C. E. Brook et al. “Optimizing COVID-19 control with asymptomatic surveillance testing in a university environment.” In: Epidemics 37 (Dec. 2021), p. 100527. issn: 1878-0067. doi: 10.1016/j.epidem.2021.100527. ppublish.

[3] “COVID-19 Stats: College and University COVID-19 Student Testing Protocols, by Mode of Instruction–United States, Spring 2021”. In: MMWR Morbidity and mortality weekly report 70(14 Apr. 2021), p. 535. issn: 1545-861X. doi: 10.15585/mmwr.mm7014a5.epublish.

[4] D. H. Hamer et al. “Assessment of a COVID-19 Control Plan on an Urban University Campus During a Second Wave of the Pandemic.” In: JAMA network open 4 (6 June 2021), e2116425. issn: 2574-3805. doi: 10.1001/jamanetworkopen.2021.16425. url: https://www.ncbi.nlm.nih.gov/pmc/articles/PMC8233704/.epublish.

[5] C. Peacock. Stanford leaders and health experts detail university’s COVID-19 surveillance testing program. Stanford University. Sept. 20, 2020. url: https://news.stanford.edu/2020/09/02/university-med-school-leaders-discuss-campus-covid-testing/ (visited on 04/24/2023).

[6] T. N. Denny et al. “Implementation of a Pooled Surveillance Testing Program for Asymptomatic SARS-CoV-2 Infections on a College Campus - Duke University, Durham, North Carolina, August 2-October 11, 2020.” In: MMWR. Morbidity and mortality weekly report 69 (46 Nov. 2020), pp. 1743–1747. issn: 1545-861X. doi: 10.15585/mmwr.mm6946e1. epublish.

[7] J. P. Bigouette et al. “Application of a Serial Antigen-Based Testing Strategy for Severe Acute Respiratory Syndrome Coronavirus 2 and Student Adherence in a University Setting: Wisconsin, October-November 2020.” In: Open forum infectious diseases 8 (10 Oct. 2021), ofab472. issn: 2328-8957. doi: 10.1093/ofid/ofab472. epublish.

[8] K. Drenkard et al. “University COVID -19 Surveillance Testing Center: Challenges and Opportunities for Schools of Nursing.” In: Journal of professional nursing : official journal of the American Association of Colleges of Nursing 37 (5 2021), pp. 948–953. issn: 1532-8481. doi: 10.1016/j.profnurs.2021.07.004. ppublish.

[9] Reopening Guidelines Committee. Considerations for Reopening Institutions ofHigher Education for the Spring Semester 2021. American College Health Association, Dec. 29, 2020. url: https://www.acha.org/documents/resources/guidelines/ACHA_Considerations_for_Reopening_IHEs_for_Spring_2021.pdf.

[10] G. Yamey and R. P. Walensky. “Covid-19: re-opening universities is high risk.” In: BMJ (Clinical research ed.) 370 (Sept. 2020), p. m3365. issn: 1756-1833. doi: 10.1136/bmj.m3365. epublish.

[11] E. Whitford. COVID-19 Mitigation Costs Still Add Up After Students Sent Home. Inside Higher Ed. Oct. 8, 2020. url: https://www.insidehighered.com/news/2020/10/09/trying-curb-covid-19-campus-expensive-whether-colleges-plans-work-or-not (visited on 06/28/2023).

[12] M. Peplow. How one university built a COVID-19 screening system. Chemical and Engineering News. Nov. 1, 2020. url: https://cen.acs.org/biological-chemistry/infectious-disease/one-university-built-COVID-19/98/i42?PageSpeed=noscript (visited on 06/28/2023).

[13] A. D. Paltiel, A. Zheng, and R. P. Walensky.”Assessment of SARS-CoV-2 Screening Strategies to Permit the Safe Reopening of College Campuses in the United States.” In: JAMA network open 3 (7 July 2020), e2016818. issn: 2574-3805. doi: 10.1001/jamanetworkopen.2020.16818. epublish.

[14] E. Losina et al. “College Campuses and COVID-19 Mitigation: Clinical and Economic Value.” In: Annals of internal medicine 174 (4 Apr. 2021), pp. 472–483. issn: 1539-3704. doi: 10.7326/M20-6558.ppublish.

[15] E. H. Bradley, M.-W. An, and E. Fox. “Reopening Colleges During the Coronavirus Disease 2019 (COVID-19) Pandemic-One Size Does Not Fit All.” In: JAMA network open 3 (7 July 2020), e2017838. issn: 2574-3805. doi: 10.1001/jamanetworkopen.2020.17838. epublish.

[16] E. Brooks-Pollock et al. High COVID-19 transmission potential associated with re-opening universities can be mitigated with layered interventions. 2021. doi: 10.1101/2020.09.10.20189696.

[17] F. C. Motta et al. “Assessment of Simulated Surveillance Testing and Quarantine in a SARS-CoV-2-Vaccinated Population of Students on a University Campus.” In: JAMA health forum 2 (10 Oct. 2021), e213035. issn: 2689-0186. doi: 10.1001/jamahealthforum.2021.3035. epublish.

[18] G. D. Lyng et al. “Identifying optimal COVID-19 testing strategies for schools and businesses: Balancing testing frequency, individual test technology, and cost.” In: PloS one 16 (3 2021), e0248783. issn: 1932-6203. doi: 10.1371/journal.pone.0248783. epublish.

[19] P. I. Frazier et al. “Modeling for COVID-19 college reopening decisions: Cornell, a case study.” In: Proceedings of the National Academy of Sciences of the United States of America 119 (2 Jan. 2022). issn: 1091-6490. doi: 10.1073/pnas.2112532119. ppublish.

[20] P. V. Markov et al. “The evolution of SARS-CoV-2.” In: Nature reviews. Microbiology (Apr. 2023). issn: 1740-1534. doi: 10.1038/s41579-023-00878-2. aheadofprint.

[21] State University of New York. Chancellor Malatras Announces Program to Begin More Easily Vaccinating SUNY Residential Students Prior to Spring Semester Departure Using One Dose Vaccine. Apr. 6, 2021. url: https://www.suny.edu/suny-news/press-releases/4-21/4-6-21/jj-vaccine-students.html (visited on 05/18/2023).

[22] Abbott. BinaxNOW: With a swab and a card, reliable coronavirus test results in 15 minutes. Aug. 26, 2020. url: https://www.abbott.com/corpnewsroom/diagnostics-testing/upping-the-ante-on-COVID-19-antigen-testing.html (visited on 05/18/2023).

[23] Cue Health. Cue COVID-19 Test Instructions For Use–For Professional Use. Tech. rep. Mar. 26, 2021. url: https://www.fda.gov/media/138826/download (visited on 05/18/2023).

[24] T. Enamorado, B. Fifield, and K. Imai. fastLink: Fast Probabilistic Record Linkage with Missing Data. R package version 0.6.0. 2020. url: https://CRAN.R-project.org/package=fastLink.

[25] R Core Team. R: A Language and Environment for Statistical Computing. R Foundation for Statistical Computing. Vienna, Austria, 2015. url: http://www.R-project.org/.

[26] A. Zeileis and C. Kleiber. countreg: Count Data Regression. R package version 0.2-1. 2022. url: https://R-Forge.R-project.org/projects/countreg/.

[27] A. Zeileis, C. Kleiber, and S. Jackman. “Regression Models for Count Data in R”. In: Journal of Statistical Software 27.8 (2008), 1–25. doi: 10.18637/jss.v027.i08. url: https://www.jstatsoft.org/index.php/jss/article/view/v027i08.

[28] C. Kleiber and A. Zeileis. “Visualizing Count Data Regressions Using Rootograms”. In: The American Statistician 70.3 (2016), pp. 296–303. doi: 10.1080/00031305.2016.1173590.

[29] K. Whtie, W. TF, and G. BG. “The ecology of medical care.” In: The New England journal of medicine 265 (Nov. 1961), pp. 885–892. issn: 0028-4793. doi: 10.1056/NEJM196111022651805. ppublish.

[30] K. J. Roghmann and R. J. Haggerty. “The diary as a research instrument in the study of health and illness behavior: experiences with a random sample of young families.” In: Medical care 10 (2 1972), pp. 143–163. issn: 0025-7079. doi: 10.1097/00005650-197203000-00004. ppublish.

[31] L. A. Green et al. “The ecology of medical care revisited.” eng. In: N Engl J Med 344.26 (2001), pp. 2021–2025. doi: 10.1056/NEJM200106283442611. url: http://dx.doi.org/10.1056/NEJM200106283442611.

[32] B.-M. Bernhardson et al. “Sensations, symptoms, and then what? Early bodily experiences prior to diagnosis of lung cancer.” In: PloS one 16 (3 2021), e0249114. issn: 1932-6203. doi: 10.1371/journal.pone.0249114. epublish.

[33] G. J. Wilde. Target risk: Dealing with the danger of death, disease and damage in everyday decisions. en. 1994. isbn: 0969912404.

[34] G. J. S. Wilde. “Risk homeostasis theory: an overview”. In: 4 (1998), pp. 89–91. issn: 1353-8047. doi: 10.1136/ip.4.2.89.

[35] M. Wanat et al. “Perceptions on undertaking regular asymptomatic self-testing for COVID-19 using lateral flow tests: a qualitative study of university students and staff.” In: BMJ open 11 (9 Sept. 2021), e053850. issn: 2044-6055. doi: 10.1136/bmjopen-2021-053850. epublish.

[36] J. A. Hirst et al. “Feasibility and Acceptability of Community Coronavirus Disease 2019 Testing Strategies (FACTS) in a University Setting.” In: Open forum infectious diseases 8 (12 Dec. 2021), ofab495. issn: 2328-8957. doi: 10.1093/ofid/ofab495. epublish.

[37] L. Bauld et al. “Students’ and staffs’ views and experiences of asymptomatic testing on a university campus during the COVID-19 pandemic in Scotland: a mixed methods study.” In: BMJ open 13 (3 Mar. 2023), e065021. issn: 2044-6055. doi: 10.1136/bmjopen-2022-065021. epublish.

[38] H. Blake et al. “Students’ Views towards Sars-Cov-2 Mass Asymptomatic Testing, Social Distancing and Self-Isolation in a University Setting during the COVID-19 Pandemic: A Qualitative Study.” In: International journal of environmental research and public health 18 (8 Apr. 2021). issn: 1660-4601. doi: 10.3390/ijerph18084182. epublish.

[39] C. E. French et al. “Low uptake of COVID-19 lateral flow testing among university students: a mixed methods evaluation.” In: Public health 204 (Mar. 2022), pp. 54–62. issn: 1476-5616. doi: 10.1016/j.puhe.2022.01.002. ppublish.

