## Supplementary material for "Assessment of the effectiveness of required weekly COVID-19 surveillance antigen testing at a university": Assessment of Model Fit

Christopher W. Ryan, MD, MS, MSPH

2023-11-22 15:15:09

### 1 Assessing fit of the zero-inflated negative binomial model

Several modeling strategies are available for count data like the number of symptomatic days prior to specimen collection that is of interest here. A common option is the Poisson model. The negative binomial model extends the Poisson to situations with overdispersion—more variation in the counts than a Poisson model could account for. A zero-inflated negative binomial model extends that further, to the context of more zero counts than the negative binomial could accomodate. Since the purpose of a surveillance testing system for COVID-19 is to detect infected people (cases) early, ideally before symptoms begin, an "excess" or "inflated" number of zero counts might be expected in the current study.

Methods to test for the presence of these "excess" zero counts in the Poisson context have been developed.[1,2] Ye, He, and colleagues have extended their test for excess zero counts to the negative binomial context.[3] Applying the He test to the present dataset yielded a test statistic of 13.7. Since, under the null hypothesis of no excess zeros, the He test statistic has a standard normal distribution, this provides strong evidence that a zero-inflation component is required to accurately model these data.

Graphical assessments of model fit yield similar evidence. In Figure 1, the rootogram in subfigure 1a shows a close match between the observed and predicted counts from the zero-inflated model, and the probability integral transform plot in subfigure 1b is close to a uniform distribution, again suggesting a good fit between data and model. By way of contrast, Figure 2 shows similar diagnostic plots from a negative binomial model without zero-inflation; it does not fit the data as well.

### References


[1] H. He et al. "A test of inflated zeros for Poisson regression models." In: *Statistical methods in medical research* 28 (4 Apr. 2019), pp. 1157–1169. ISSN: 1477-0334. DOI: 10.1177/0962280217749991. ppublish.

[2] Y. Tang and W. Tang. "Testing modified zeros for Poisson regression models." In: *Statistical methods in medical research* 28 (10-11 2019), pp. 3123–3141. ISSN: 1477-0334. DOI: 10.1177/0962280218796253. ppublish.

[3] P. Ye et al. "Testing latent class of subjects with structural zeros in negative binomial models with applications to gut microbiome data." In: *Statistical methods in medical research* 31 (11 Nov. 2022), pp. 2237–2254. ISSN: 1477-0334. DOI: 10.1177/09622802221115881. ppublish.

Figure 1: Diagnostic plots for the zero-inflated negative binomial model

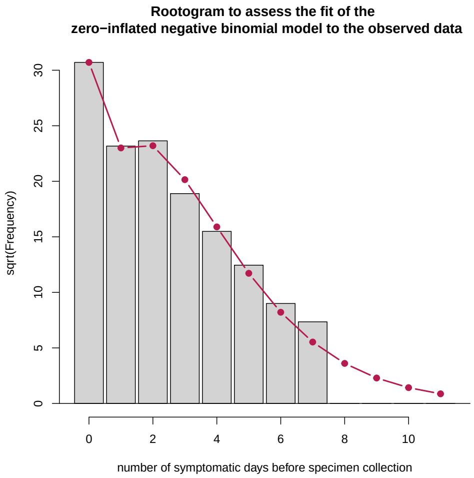


(a) Rootogram.The grey bars represent observed counts of subjects with the number of symptomatic days (shown on the horizontal axis) prior to specimen collection. The red line shows the model's predictions. Observed counts beyond 7 days are not shown.

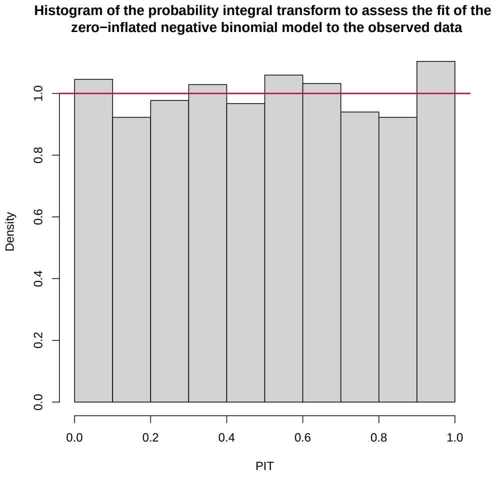


(b) Probability integral transform histogram. The more uniform the height of the grey bars, the better the fit of the model to the observed data.

Figure 2: Diagnostic plots for a negative binomial model without zero-inflation

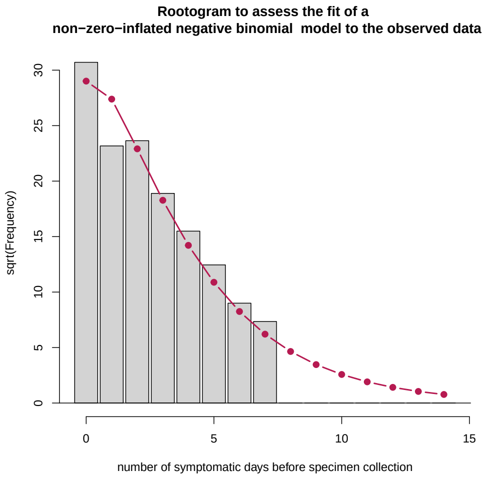


(a) Rootogram.The grey bars represent observed counts of subjects with the number of symptomatic days (shown on the horizontal axis) prior to specimen collection. The red line shows the model's predictions. Observed counts beyond 7 days are not shown.

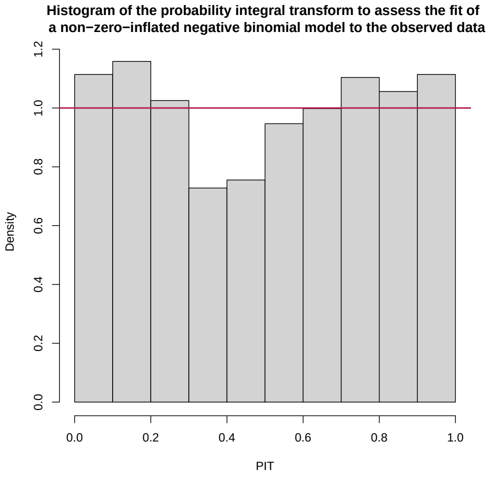


(b) Probability integral transform histogram. The more uniform the height of the grey bars, the better the fit of the model to the data.
